# Modulation of Posterior Insula Selectively Enhances Nociceptive Sensory Gating in Humans

**DOI:** 10.64898/2026.01.16.26344114

**Authors:** Gabriel Isaac, Aditya Kapoor, Andrew Strohman, Wynn Legon

**Affiliations:** School of Neuroscience, Virginia Polytechnic Institute and State University, Blacksburg, VA, 24061, USA; Fralin Biomedical Research Institute at Virginia Tech Carilion, Roanoke, VA, 24016, USA; Virginia Tech Carilion School of Medicine, Roanoke, VA, 24016, USA; Graduate Program in Translational Biology, Medicine, and Health, Virginia Polytechnic Institute and State University, Roanoke, VA, 24016, USA; Center for Human Neuroscience Research, Fralin Biomedical Research Institute at Virginia Tech Carilion, Roanoke, VA, 24016, USA; Center for Health Behaviors Research, Fralin Biomedical Research Institute at Virginia Tech Carilion, Roanoke, VA, 24016, USA; Department of Neurosurgery, Carillion Clinic, Roanoke, VA, 24016, USA

## Abstract

Sensory gating — the brain’s ability to filter out repetitive sensory input — is essential for preventing sensory overload. Impaired gating is frequently observed in nociplastic and other chronic overlapping pain conditions, yet the specific brain regions supporting this inhibitory process in humans remains unclear. Neuroimaging studies examining pain processing implicate the anterior insula (AI), posterior insula (PI), and anterior mid-cingulate cortex (aMCC), but their deeper locations limit direct mechanistic testing using conventional non-invasive techniques. Here, we leveraged low-intensity focused ultrasound (LIFU), a novel non-invasive neuromodulation method with high depth-penetration and millimeter resolution, to examine the contributions of the AI, PI, and aMCC to sensory gating of nociceptive stimuli. Twelve healthy adults completed four counterbalanced visits of a paired-pulse contact heat evoked potential (CHEP) paradigm while receiving LIFU targeted to each region or an active sham. Using surface electroencephalography (EEG), placed at site Cz, we quantified the peak-to-peak (P2P) amplitude of the cortical response to the first stimulus (S1), the second stimulus (S2), and used the ratio of the response to each stimulus (S2/S1 ratio) as an index of sensory gating. Subjective ratings of pain intensity to the second stimulus were also recorded. Results demonstrated that all subjects displayed sensory gating at baseline and thatLIFU produced region-specific effects. Both PI and aMCC neuromodulation reduced subjective pain ratings and significantly decreased S2 amplitude relative to sham, whereas LIFU to AI had no effect. Critically, only PI neuromodulation enhanced sensory gating by reducing the S2/S1 ratio. These findings identify the PI as a key contributor to gating of repetitive nociceptive input and a promising neuromodulation target for remediating sensory gating deficits in nociplastic pain.

## Introduction

The brain’s ability to filter redundant nociceptive and non-painful sensory input is a fundamental physiological mechanism of adaptation and characteristic of healthy sensory systems [1,2]. This process, broadly described as sensory gating or habituation [3], reflects inhibitory control mechanisms that dampen ascending sensory input resulting from repetitive stimulation across modalities [4–7]. Disruption of this inhibition can lead to sensory over-responsiveness, hypervigilance, and maladaptive pain perception [8,9]. Dysfunctional sensory gating has been proposed as a hallmark of many nociplastic and chronic overlapping pain conditions which are characterized by hypersensitivity to both painful and innocuous sensory stimulation [10–12]. Clarifying the brain regions coordinating sensory gating could provide a better foundation to examine the disrupted nociceptive processing observed in patients with chronic pain.

One method to investigate sensory gating mechanisms is through paired pulse evoked potentials recorded using surface electroencephalography (EEG). Paired-pulse evoked potential paradigms involve presenting pairs of identical stimuli in quick succession and comparing the magnitude of the evoked response of the second stimulus (S2) relative to the first (S1) [5,13–15]. In healthy sensory systems, the response to S2 is reduced (S2/S1 < 1), demonstrating both inhibition of repetitive sensory input [16,17], and down-regulation of stimulus saliency [18]. Diminished attenuation or even facilitation of the S2 response (S2/S1 ≥ 1) suggests impaired inhibitory mechanisms and/or sensory hypervigilance and has been observed as a distinguishing feature in nociplastic pain conditions [9,11,19].

The contact heat evoked potential (CHEP) is a prominent biphasic wave (N2-P2) recorded maximally at the scalp vertex (Cz) via surface EEG, typically peaking at ∼ 500 milliseconds after stimulation [20,21]. It is a common measure of the brain response to a nociceptive stimulus that may reflect a combination of both stimulus intensity [22] and salience attribution [23,24], an additional process known to be dysfunctional in nociplastic and chronic overlapping pain [25].

Multiple lines of evidence consistently implicate the insula and anterior mid-cingulate cortex (aMCC) as key hubs coordinating the inhibitory control and salience appraisal of ascending nociceptive input [26–31]. The insula can be divided into the anterior insula (AI) and the posterior insula (PI), subregions which have been shown to play distinct roles in nociceptive processing [32–34]. The PI receives afferent nociceptive input via the ventromedial posterior thalamus and is thought to encode qualities of pain such as modality, intensity and frequency [35–37]. This information is then transferred to the AI which is more heavily interconnected with higher-order affect and salience regions such as the amygdala and aMCC and is thought to assign nociceptive input its behavioral relevance and emotional significance [38,39]. Together, these regions form a posterior-to-anterior gradient responsible for the appraisal of ascending nociceptive signals [39]. The aMCC serves a complementary role, integrating direct and early nociceptive input via the medial dorsal nucleus of the thalamus with motivational, salience, and cognitive signals thought to support action selection [40–42]. Through their coordinated activity, the insula and aMCC look to filter and shape nociceptive information and the subjective experience of pain to inform decision making [43]. In nociplastic pain conditions, hyperactivity of both the insula and aMCC in response to painful and non-painful stimuli is consistently reported [44–48]. This aberrant activity may reflect impaired sensory gating and over-appraisal of stimulus saliency [30], though the causal contributions of insular subregions and the aMCC to innocuous sensory and nociceptive processing remain unclear.

The insula and the aMCC are deep to the scalp surface (∼ 4 - 5 cm) precluding intervention using conventional non-invasive neuromodulation tools such as transcranial magnetic or electrical stimulation [49,50]. Low-intensity focused ultrasound (LIFU) is a promising alternative to these conventional methods, enabling safe, noninvasive modulation of cortical and subcortical structures with millimeter precision [51,52]. Human studies demonstrate that LIFU can modulate cortical excitability [51,53] and reduce pain-evoked brain responses [54,55], establishing it as a versatile tool for causally probing the neural mechanisms that govern sensory gating in the human brain.

Given the evidence implicating the AI, PI, and aMCC in inhibitory control and pain processing [26–31], we sought to test whether LIFU targeted to these regions modulates cortical sensory gating in healthy adults. Participants completed two blocks of a paired-pulse paradigm: a baseline block involving pairs of identical contact heat stimuli, followed by an intervention block in which the same paradigm was repeated during concurrent LIFU neuromodulation to the AI, PI, aMCC, or an active sham condition. Pairs of CHEPs (S1 and S2) were recorded and the S2/S1 amplitude ratio was calculated to index sensory gating, alongside subjective ratings of stimulus intensity to S2. Given that pain-evoked potentials index aspects of stimulus salience [56], we predicted that targeting salience-network nodes (AI, aMCC) would have the greatest effect on sensory gating, resulting in lower S2/S1 ratios compared with PI or sham neuromodulation. Additionally, we hypothesized that LIFU to the aMCC and PI would significantly reduce subjective pain ratings relative to sham neuromodulation given their early sensory processing of nociceptive input and their observed roles in specifically encoding of features of pain such as intensity [57].

## MATERIALS & METHODS

### Participants

All experimental procedures were approved by the Virginia Tech Institutional Review Board (VT-IRB #21-796) and the study was registered on ClinicalTrials.gov (NCT05145426). A total of 12 healthy volunteers (mean age: 33 ± 13 years; range = 21-61 years; 8 female, 4 male) provided written informed consent and received financial compensation for their participation. Exclusion criteria included current or chronic pain, contraindications to noninvasive neuromodulation (as outlined by Rossi et al. for transcranial magnetic stimulation) [58], contraindications to MRI or CT (including pregnancy), active medical conditions or treatments with potential central nervous system effects (e.g., Alzheimer’s disease), a history of neurological disorders (e.g., Parkinson’s disease, epilepsy, essential tremor), prior head injury with loss of consciousness >10 minutes, or a history of alcohol or drug dependence.

### Overall Study design

This study used a single-blind, active sham-controlled crossover design conducted over five visits on separate days, with a minimum interval of two days between visits. In the first visit, participants underwent anatomical MRI and CT scans (used for targeting and acoustic modelling – see below) as well as baseline assessments. In visits 2 through 5, participants received LIFU targeted to one of three brain targets: the AI, PI, or aMCC, or an active Sham condition. Intervention order was randomized and counterbalanced across participants, with the Sham condition randomly assigned to either the AI, PI, or aMCC site (**Figure 1A**).

**Figure 1.**
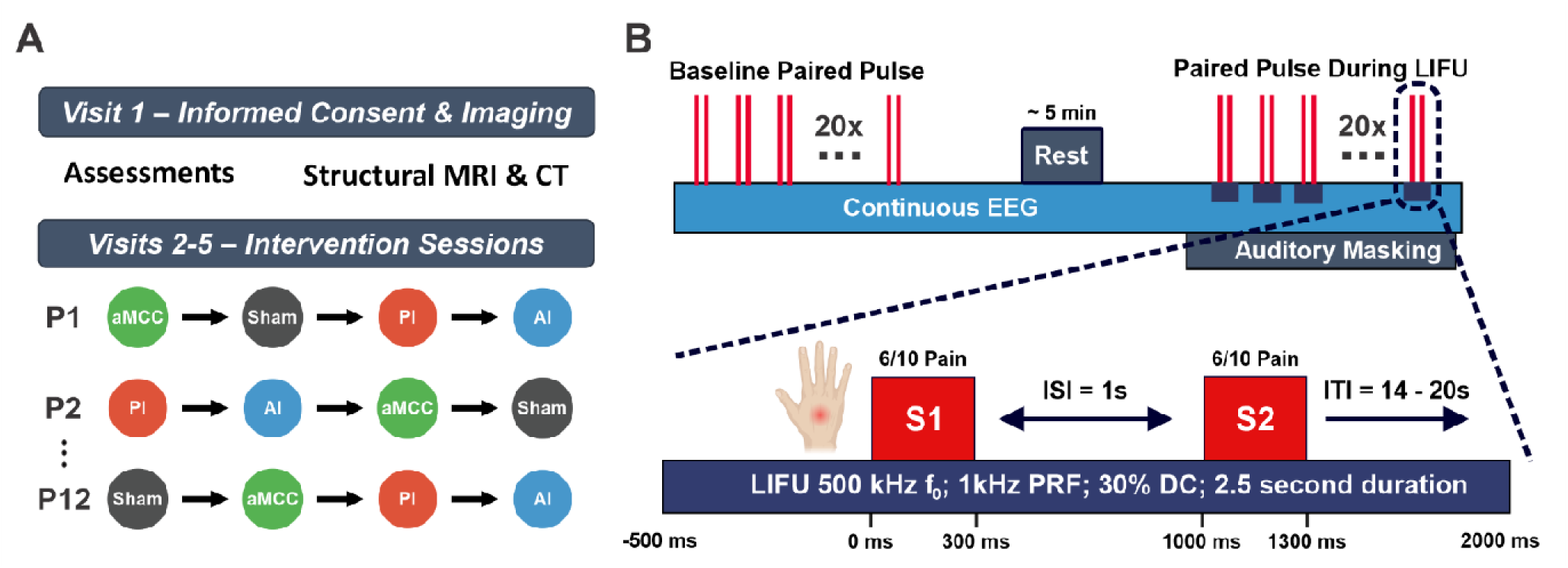
Experimental design and task timing. **A.** 12 healthy participants (labelled P1 through P12) completed five visits. Visit 1 involved magnetic resonance imaging (MRI) and computed tomography (CT) imaging as well as baseline assessments. Visits 2-5 were single-blind, sham-controlled intervention visits delivering LIFU to the anterior insula (AI), posterior insula (PI), anterior midcingulate (aMCC), or an active Sham in a randomized, counterbalanced order. **B.** (Top) Participants completed a paired-pulse task during two periods: an initial baseline block and a second block delivered during LIFU neuromodulation. Each block consisted of 20 pairs of contact heat stimuli (vertical red lines) during continuous EEG recording. A brief rest period separated the two blocks, and auditory masking was used during the LIFU block to blind participants. (Bottom) Each trial consisted of two identical heat stimuli delivered to the dorsum of the right hand (S1 and S2), each lasting 300 ms (50-ms ramp up, 200-ms plateau, 50-ms ramp down). Stimulus intensity was individually adjusted to produce moderate pain (6/10). The second stimulus (S2) occurred 1 s after the first (inter-stimulus interval, ISI), and trials were separated by a jittered 14-20 s inter-train interval (ITI). LIFU (parameters: 500 kHz, 1 kHz pulse repetition frequency, 30% duty cycle) delivery was time-locked to the first heat stimulus, beginning 500 ms before S1 and continuing for a total of 2.5 s.

### Assessments

On each testing day (Visits 2-5), participants completed assessments immediately before and after LIFU. Pre-visit assessments included a Report of Symptoms (ROS) [59]. The ROS documented the presence and severity (absent, mild, moderate, severe) of symptoms such as headache and sleepiness before and after LIFU following the approach detailed in Legon et al. (2020) [59]. An Experience Questionnaire [54,55], queried whether participants could hear or feel LIFU and whether they believed they received active stimulation. Items included: “I could hear the LIFU stimulation,” “I could feel the LIFU stimulation,” and “I believe I experienced LIFU stimulation.” Responses were collected on a 7-point Likert scale (0 = strongly disagree, 3 = neutral, 6 = strongly agree).

### Control Measures

#### Blinding

Both participants and the data analysis team were blinded to condition assignment. The allocation sequence was stored separately and not revealed until all analyses were complete. The experimenters administering stimulation were aware of the assignment but were not involved in data analysis. While this minimized risk of bias, it remained possible that researchers could have inadvertently conveyed condition allocation to participants.

#### Active Sham Stimulation

Sham stimulation was delivered using the same transducer and coupling procedures (see “LIFU Transducers and Waveform” below) as active visits, but the ultrasound beam was blocked by an acoustically opaque attenuator [60], allowing participants to experience the same auditory and procedural cues without sonication. Intervention order was pseudorandomized and counterbalanced across participants, with the Sham condition assigned to either the AI, PI, or aMCC site.

#### Auditory Masking

In some cases, LIFU transducers can produce audible artifacts likely due to the pulse repetition frequency occurring within the human range of hearing. To eliminate this potential confound, strict acoustic masking was implemented. Participants wore disposable earbuds connected to a Kindle tablet running a commercially available white noise application (*White Noise Baby Sleep Sounds*, AMICOOLSOFT). A multitone noise, previously demonstrated to effectively mask ultrasound artifacts [61], was selected within the app and applied across all participants. The volume was set to a comfortable level that also minimized ambient sounds, typically averaging 70-75 dB. Masking was verified by speaking to participants outside their visual field. The noise was played continuously throughout the testing visit. Thirty minutes after testing, participants completed the Experience Questionnaire.

### Paired Pulse delivery and recording

#### Paired Pulse paradigm

Paired pulse stimuli were generated using a contact heat stimulator (QST.Lab, Strasbourg, France) applied to the dorsum of the right hand. Each heat pulse was 300 ms (50-ms rise/fall, 200-ms plateau; baseline 32 °C). A thresholding procedure was first completed to standardize experimental pain across participants. Three heat stimuli were delivered, each separated by 5 s, and participants rated the subjective pain intensity. The temperature was then increased by 1 °C and the procedure repeated until a pain rating of 6/10 was achieved or the stimulator reached its maximum temperature (60 °C). Stimulation sites were distributed across a 2 x 3 grid spaced about 1 inch apart to minimize potential skin irritation that may confound habituation mechanisms. Prior to administering LIFU, participants completed a baseline paired-pulse block. This block was used to account for potential day-to-day variability in evoked-potential amplitude but also to identify non-responders whose evoked potentials could not be reliably measured. This block consisted of 20 pairs of identical heat stimuli separated by 1 s delivered at the threshold intensity, with inter-train intervals (ITIs) jittered between 14-20 s (**Figure 1B**). Both the first (S1) and second (S2) evoked potentials were required to be clearly visible in the EEG recording. If either was not detectable, the electrodes were re-prepared and testing was repeated. If either response remained absent after re-preparation, the visit was cancelled and rescheduled. Following a successful baseline block, the same 20-pair paired-pulse CHEP paradigm was repeated concurrent with LIFU neuromodulation (see LIFU transducer section below for details). During both baseline and LIFU blocks, participants rated their subjective pain intensity of the S2 stimulus using a 0-10 numerical rating scale where 0 = “no pain” and 10 = “worst pain imaginable” [62]. Pain ratings were entered using a keypad operated with the left hand and were recorded using a custom MATLAB script.

#### Electroencephalography (EEG) recording

EEG was recorded using an EEG amplifier module (EEG100C) interfaced with a BIOPAC MP160 data acquisition system (BIOPAC Systems Inc., Goleta, CA) and a 10-mm silver-silver chloride cup electrode placed at the vertex (Cz) referenced to the right mastoid. A ground electrode was placed at the right elbow. The scalp was prepared with a mild abrasive gel (Nuprep; Weaver and Company, Aurora, CO) followed by rubbing alcohol. Electrodes were filled with conductive paste (Ten20 Conductive; Weaver and Company) and secured with medical tape. Data were sampled at 1000 Hz, with the electrode impedance maintained below 50 kΩ throughout the visit.

#### EEG processing

EEG data were band-pass filtered from 2–100 Hz using a third-order Butterworth filter with zero-phase shift using the `filtfilt` function in MATLAB. Data were visually inspected for ocular, muscle, or motion-related artifacts, and contaminated trials were removed. Across participants, no more than three of the twenty paired-pulse trials were rejected, with an average of 1 rejection per participant. Clean trials were epoched from -500 to 2500 ms relative to S1 onset and baseline-corrected using the -1500 to -500 ms pre-S1 interval. Trials were then averaged within each block (baseline and LIFU) to generate mean S1 and S2 waveforms. N2-P2 P2P amplitudes and latencies for S1 and S2 were extracted using a custom manual peak-detection script requiring a distinct inflection for component inclusion.

### Low-intensity focused ultrasound (LIFU)

#### LIFU transducers and waveform

Two different single-element focused ultrasound transducers were used depending on the target. For the insula (AI, PI) we used an H-281 (500 kHz center frequency; active diameter 45.0 mm; geometric focus 45.0 mm; focal depth from exit plane 38.0 mm) (Sonic Concepts, Bothell, WA). For aMCC we used a H-104 (500 kHz center frequency; aperture 64 mm; focal length 52 mm from the exit plane) (Sonic Concepts, Bothell, WA). Transducers were coupled to the scalp with ultrasound gel and individualized stand-off pucks [63] when needed to help align the geometric focus to target depth. During sonication, the transducer was manually held against the scalp by a trained researcher and guided by a neuronavigation system (BrainSight; Rogue Research, Montreal, QC, Canada), to maintain accurate and stable targeting throughout the experiment.

The waveform was generated via a dual-channel function generator (4078B, BK Precision). Channel 2 produced a 500 kHz sine carrier. Channel 1 generated a 1 kHz gating signal that pulsed the carrier to achieve a 30% duty cycle. The output was amplified with a 100 W RF amplifier (E&I 2100L; Electronics & Innovation) and delivered through the transducer’s matching network. Each trial consisted of a 2.5 s pulse train (N = 2500 pulses at 1 kHz PRF; pulse width = 300 μs; duty cycle = 30%), time-locked to begin 500 ms prior to the S1 heat stimulus onset and end 2 seconds afterwards so that LIFU was on during the generation of both S1 and S2 (**Figure 1B**). Sonication parameters were chosen based upon prior work that systematically tested a range of parameter sets, with the present study adopting the set that yielded the most reliable inhibition [64].

### Acoustic Field Mapping and Acoustic/Thermal Modeling

#### Empirical acoustic testing

Pressure fields were measured in a degassed water tank using a calibrated hydrophone (HNR-0500, Onda Corp.) mounted on a motorized 3D stage. XY and YZ planar scans were collected at 0.25 mm resolution to define the focal spot and full-width at half-maximum (FWHM) dimensions. Input-output sweeps (20-250 mVpp) established the relationship between driving voltage and acoustic pressure at the focus. These measurements were used to calibrate stimulation parameters for in vivo visits where the applied driving voltage was standardized to deliver the same extracranial pressure across all participants. Normalized pressure maps are visualized in **Figure S1A**.

#### Acoustic modelling

Acoustic simulations were run using each participant’s MR and CT images to evaluate LIFU wave propagation through the skull and to estimate intracranial acoustic pressure distributions (**Figure 2**). Simulations were conducted using the k-Wave MATLAB toolbox [65], which implements a pseudospectral time-domain method to solve discretized acoustic wave equations on a spatial grid. CT images were used to construct the acoustic model of the skull, whereas MR images were used to define LIFU targets within brain based on individual anatomy. Modeling parameters and procedures followed those reported in prior work [66]. The computational model of the ultrasound transducer was designed to replicate empirical pressure maps measured in a water tank, consistent with previous studies [54,55].

**Figure 2.**
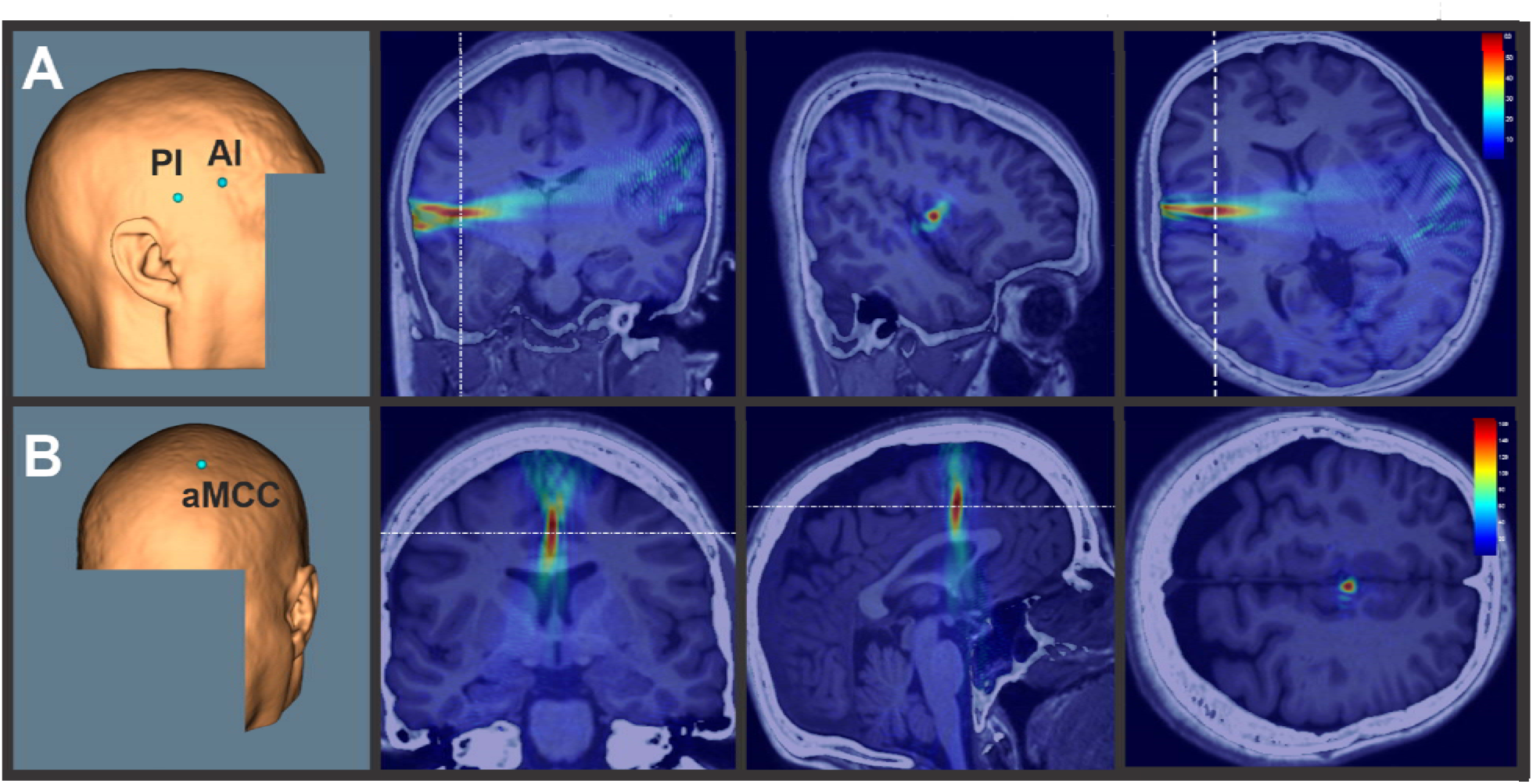
Scalp targets & Acoustic models. **A.** (Left) Scalp targets taken from the neuronavigation system showing placement of the transducer center for the anterior insula (AI) and posterior insula (PI) from one representative subject. (Right) Single participant acoustic modeling based on individual magnetic resonance (MR) and computed tomography (CT) scans illustrating targeting of the PI in coronal, sagittal, and transverse views. Transverse view is taken from dashed white line in coronal and sagittal view. The color bar indicates acoustic pressure in kilopascals (kPa). **B.** (Left) Scalp target for the anterior mid-cingulate cortex (aMCC) from one representative participant. (Right) Single participant acoustic modeling illustrating targeting of the aMCC in the coronal, sagittal, and transverse planes. Transverse view is taken from dashed white line in coronal and sagittal view. Color bar indicates acoustic pressure in kPa.

#### Thermal Modelling

Thermal simulations were performed using the modified mixed-domain method (mSOUND) with a three-layer skull model (cortical-trabecular-cortical) based on Benchmark 6 from Aubry et al. (2022) [67]. We modeled a single parameter set corresponding to a conservative, higher-than-experimental extracranial dose: 30% duty cycle, 2.5-s pulse duration, 10 Hz PRF, 750 kPa outside the skull. For computational feasibility, the simulation was run at 10 Hz PRF instead of 1 kHz; prior work has shown that lowering PRF while holding total energy constant produces negligible differences in predicted thermal rise [68]. Stimulation consisted of 20 repetitions, each separated by a 14-s interstimulus interval-the minimum ISI used in this study. To provide a conservative estimate of heating, the simulation assumed a 65% absorption through the skull, with all absorbed energy converted to heat (**Figure S1B**).

### Outcome Measures and Statistical Analysis

#### Assessments

Responses from the auditory masking assessments (7-point Likert scale; 0-6) were analyzed using nonparametric Kruskal-Wallis tests across conditions. Scores from the experience questionnaire are summarized as mean ± SD.

#### Pain ratings

For each participant, subjective pain intensity scores from the twenty S2 stimuli were averaged for baseline and each LIFU block. Baseline ratings were compared across stimulation sites using a one-way repeated-measures ANOVA to test for day-to-day differences prior to the LIFU intervention. To assess LIFU effects, difference scores were calculated for each participant by subtracting the average baseline rating from average LIFU rating from each condition (AI, PI, aMCC, Sham) to control for potential day-to-day differences. These difference scores were then analyzed with a one-way repeated-measures ANOVA. Significant main effects were followed up with Tukey-Kramer post hoc tests correcting for multiple comparisons. Statistical significance was defined as p < 0.05.

#### CHEP Amplitude

For both the baseline and the intervention blocks, N2-P2 peak-to-peak (P2P) amplitudes for S1 and S2 were computed from the averaged waveforms, and the S2/S1 sensory gating index was calculated (see **Figure 3A** for example paired-pulse CHEP waveform). Intervention P2P amplitudes were normalized within visit to account for day-to-day variability by dividing the S1 and S2 intervention P2P amplitudes by the same-day baseline S1 amplitude.

**Figure 3.**
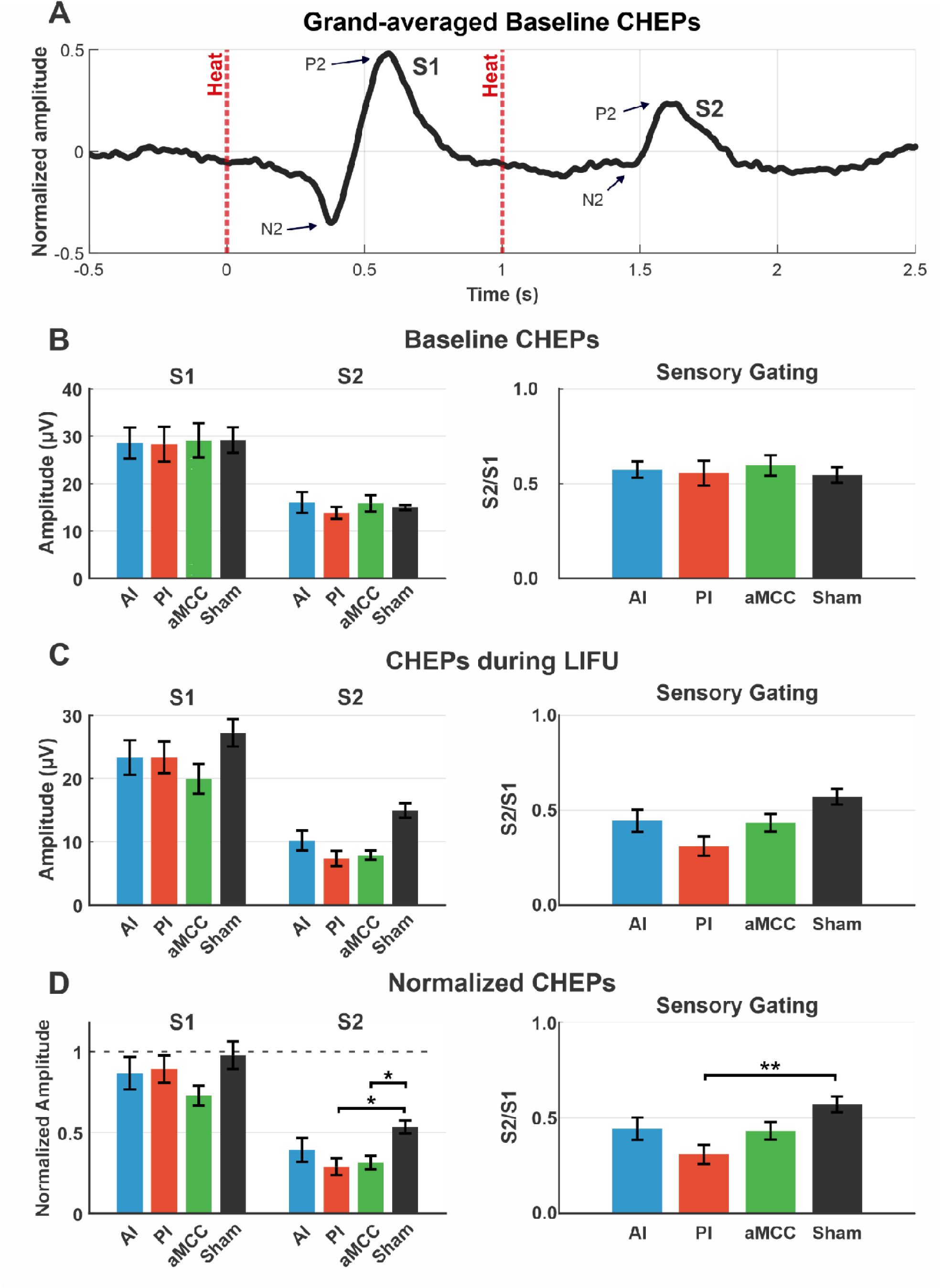
Effect of LIFU on contact heat-evoked potentials (CHEPs). **A**. Grand-averaged (N = 12) normalized baseline CHEP waveform (prior to LIFU across all visits) showing the CHEP response to the first (S1) contact heat stimulus (vertical dashed red line) and the second (S2) contact heat stimulus. CHEP amplitude was measured as peak-to-peak amplitude from the N2 to the P2 potential. **B.** (Left) Group (N = 12) baseline mean ± SEM CHEP S1 and S2 amplitudes and sensory gating (S2/S1) for AI, PI, aMCC, and Sham. No differences were observed across conditions. **C.** Group (N = 12) LIFU mean ± SEM CHEP S1 and S2 amplitudes and sensory gating. **D.** Group (N = 12) normalized mean ± SEM CHEP amplitudes during LIFU (expressed relative to each subject’s baseline). Normalized S1 amplitudes did not differ by condition. Normalized S2 amplitudes differed across conditions, with reduced amplitudes for PI (*p = 0.013) and aMCC (*p = 0.030) compared with Sham. Normalized sensory gating (S2/S1) also differed across conditions, with stronger gating for PI relative to Sham (**p = 0.003). Asterisks denote significant post hoc comparisons.

Baseline S1 and S2 amplitudes, and S2/S1 ratios were compared across stimulation sites using one-way repeated-measures ANOVAs to confirm that any observed effects reflected LIFU neuromodulation rather than group differences in the baseline data. Normalized S1 and S2 amplitudes, and the S2/S1 gating index were then analyzed using separate one-way repeated-measures ANOVAs. To control for multiple comparisons across these six ANOVA tests, p-values were corrected using the Benjamini-Hochberg (BH) false discovery rate procedure (q = 0.05), with Tukey-Kramer post hoc tests conducted on significant effects.

## RESULTS

### Contact heat-evoked potentials

#### Raw Baseline CHEP amplitude

Group N = 12 baseline S1 amplitudes (mean ± SEM) for AI, PI, aMCC, and Sham were: 28.5 ± 3.3 µV, 28.3 ± 3.7 µV, 29.1 ± 3.6 µV, and 29.2 ± 2.7 µV, respectively (**Figure 3A & B; Table 1**). To ensure no day-to-day differences in baseline values we statistically tested this. The one-way repeated measures ANOVA showed no main effect of condition: F(3,33) = 0.017, p = 0.997, η²1Z = 0.001 (BH-q = 0.997). Group baseline S2 amplitudes were 16.0 ± 2.2 µV, 13.8 ± 1.2 µV, 15.8 ± 1.7 µV, and 15.0 ± 0.5 µV. The one-way repeated measures ANOVA showed no main effect of condition: F(3,33) = 0.406, p = 0.750, η²1Z = 0.027 (BH-q = 0.997). Group baseline sensory gating (S2/S1 proportion; mean ± SEM) 0.57 ± 0.04, 0.56 ± 0.07, 0.60 ± 0.05, and 0.55 ± 0.04 for AI, PI, aMCC, and Sham, respectively. The one-way repeated measures ANOVA showed no effect of condition: F(3,33) = 0.183, p = 0.9076, η²1Z = 0.012 (BH-q = 0.997).

**Table 1.** Raw CHEP amplitudes and sensory gating before and during LIFU. Group (N = 12) mean ± SEM S1 and S2 amplitudes (µV) and sensory gating ratios (S2/S1) are shown for each condition (AI, PI, aMCC, and Sham) at baseline and during LIFU.

| Condition | Block | S1 (μV) | S2 (μV) | S2/S1 |
| --- | --- | --- | --- | --- |
| AI | Baseline | 28.5 ± 3.3 | 16.0 ± 2.2 | 0.57 ± 0.04 |
| | Intervention | 23.3 ± 2.7 | 10.2 ± 1.6 | 0.44 ± 0.06 |
| PI | Baseline | 28.3 ± 3.7 | 13.8 ± 1.2 | 0.56 ± 0.07 |
| | Intervention | 23.3 ± 2.5 | 7.3 ± 1.2 | 0.31 ± 0.05 |
| aMCC | Baseline | 29.1 ± 3.6 | 15.8 ± 1.7 | 0.60 ± 0.05 |
| | Intervention | 19.9 ± 2.3 | 7.9 ± 0.7 | 0.43 ± 0.05 |
| Sham | Baseline | 29.2 ± 2.7 | 15.0 ± 0.5 | 0.55 ± 0.04 |
| | Intervention | 27.2 ± 2.2 | 14.9 ± 1.2 | 0.57 ± 0.04 |

#### Baseline CHEP latency

N2 latencies were 0.42 ± 0.06 s, 0.41 ± 0.05 s, 0.41 ± 0.04 s, and 0.41 ± 0.05 s for AI, PI, aMCC, and Sham. The one way repeated measures ANOVA showed no effect of condition: F(3,33) = 0.13, p = 0.93. P2 latencies were 0.58 ± 0.06 s, 0.59 ± 0.05 s, 0.57 ± 0.05 s, and 0.60 ± 0.07 s. The one way repeated measures ANOVA showed no effect of condition: F(3,33) = 0.36, p = 0.79.

#### Raw LIFU CHEP Amplitude

During LIFU, S1 amplitudes (mean ± SEM) were 23.3 ± 2.7 µV, 23.3 ± 2.5 µV, 19.9 ± 2.3 µV, and 27.2 ± 2.2 µV for AI, PI, aMCC, and Sham, respectively. S2 amplitudes were 10.2 ± 1.6 µV, 7.3 ± 1.2 µV, 7.9 ± 0.7 µV, and 14.9 ± 1.2 µV. Sensory gating values (S2/S1 proportion; mean ± SEM) were: 0.44 ± 0.06, 0.31 ± 0.05, 0.43 ± 0.05, and 0.57 ± 0.04 for AI, PI, aMCC, and Sham, respectively (see **Figure 3C** and **Table 1**).

#### Normalized CHEPs During LIFU

Group normalized S1 amplitudes (divided by baseline S1 amplitude; mean ± SEM) were 0.87 ± 0.10, 0.89 ± 0.08, 0.73 ± 0.06, and 0.98 ± 0.09 for AI, PI, aMCC, and Sham, respectively. A one-way repeated measures ANOVA revealed no main effect of LIFU condition: F(3,33) = 1.508, p = 0.226, η²1Z = 0.093 (BH-q = 0.451) (**Figure 3D**). Normalized S2 amplitudes (divided by baseline S1 amplitude) were 0.39 ± 0.07, 0.29 ± 0.05, 0.31 ± 0.04, and 0.53 ± 0.04. The one-way repeated measures ANOVA revealed an effect of condition: F(3,33) = 4.166, p = 0.011, η²1Z = 0.221 (BH-q = 0.0333). Post hoc tests showed reduced normalized S2 amplitudes for PI as compared to Sham (mean difference = -0.245, p = 0.013) and aMCC compared to Sham (mean difference = -0.220, p = 0.030) (**Figure 3D & Table 2**).

**Table 2.** Normalized CHEP amplitudes and sensory gating during LIFU. Group (N = 12) mean ± SEM normalized S1 and S2 amplitudes (% of baseline) and sensory gating ratios (S2/S1) during LIFU for each condition (AI, PI, aMCC, and Sham).

| Condition | S1 | S2 | S2/S1 |
| --- | --- | --- | --- |
| **AI** | 0.87 ± 0.10 | 0.39 ± 0.07 | 0.45 ± 0.07 |
| **PI** | 0.89 ± 0.08 | 0.29 ± 0.05 | 0.29 ± 0.05 |
| **aMCC** | 0.73 ± 0.06 | 0.31 ± 0.04 | 0.43 ± 0.04 |
| **Sham** | 0.98 ± 0.09 | 0.53 ± 0.04 | 0.57 ± 0.04 |

The normalized sensory gating values (S2/S1 proportion; mean ± SEM) were 0.45 ± 0.07, 0.29 ± 0.05, 0.43 ± 0.04, and 0.57 ± 0.04 for AI, PI, aMCC, and Sham, respectively. The one way repeated measures ANOVA showed an effect of condition: F(3,33) = 4.652, p = 0.007, η²1Z = 0.241 (BH-q = 0.0333). Tukey’s post-hoc test revealed significant differences between PI and Sham (mean difference = -28%, p = 0.003) (**Figure 3D & Table 2**). No other pairwise comparisons were significant.

#### CHEP latencies during LIFU

N2 latencies were 0.37 ± 0.07 s, 0.39 ± 0.07 s, 0.38 ± 0.05 s, and 0.40 ± 0.05 s. A one-way repeated measures ANOVA revealed no main effect of LIFU condition: F(3,33) = 0.39, p = 0.76. P2 latencies were 0.57 ± 0.09 s, 0.57 ± 0.08 s, 0.56 ± 0.06 s, and 0.59 ± 0.05 s. A one-way repeated measures ANOVA revealed no main effect of LIFU condition: F(3,33) = 0.32, p = 0.82.

### Pain ratings

The mean ± SD temperature required to achieve a subjective report of 6/10 for the AI, PI, aMCC, and Sham conditions was: 59.6 ± 1.0 °C, 59.8 ± 0.6 °C, 59.7 ± 0.8 °C, and 59.3 ± 1.1 °C, respectively. A one-way repeated-measures ANOVA demonstrated no significant differences across conditions (F(3,33) = 1.13, p = 0.350, η²1Z = 0.093). At baseline, group mean ± SEM pain ratings for AI, PI, aMCC, and Sham were 3.34 ± 0.40, 3.42 ± 0.40, 3.68 ± 0.35, and 3.62 ± 0.43, respectively (**Figure 4A**). A one-way repeated-measures ANOVA indicated no significant main effect of condition: (F(3,33) = 0.92, p = 0.410, η²1Z = 0.077). During LIFU, group mean ± SEM pain ratings for AI, PI, aMCC, and Sham were 3.26 ± 0.48, 2.99 ± 0.36, 3.24 ± 0.41, and 3.83 ± 0.39, respectively (**Figure 4B**). Pain ratings were then normalized to each participant’s same-day baseline to account for potential day-to-day variability. The resulting difference scores (mean ± SEM) for AI, PI, aMCC, and Sham were -0.08 ± 0.19, -0.43 ± 0.13, -0.44 ± 0.10, and 0.21 ± 0.10, respectively (**Figure 4C**). A one-way repeated-measures ANOVA on these normalized values revealed a significant main effect of condition: (F(3,33) = 5.66, p = 0.008, η²1Z = 0.340). Post-hoc Tukey-Kramer tests revealed that LIFU to both PI and aMCC significantly reduced pain intensity ratings relative to Sham (PI vs. Sham: p = 0.001; aMCC vs. Sham: p = 0.007). No other pairwise comparisons reached significance.

**Figure 4.**
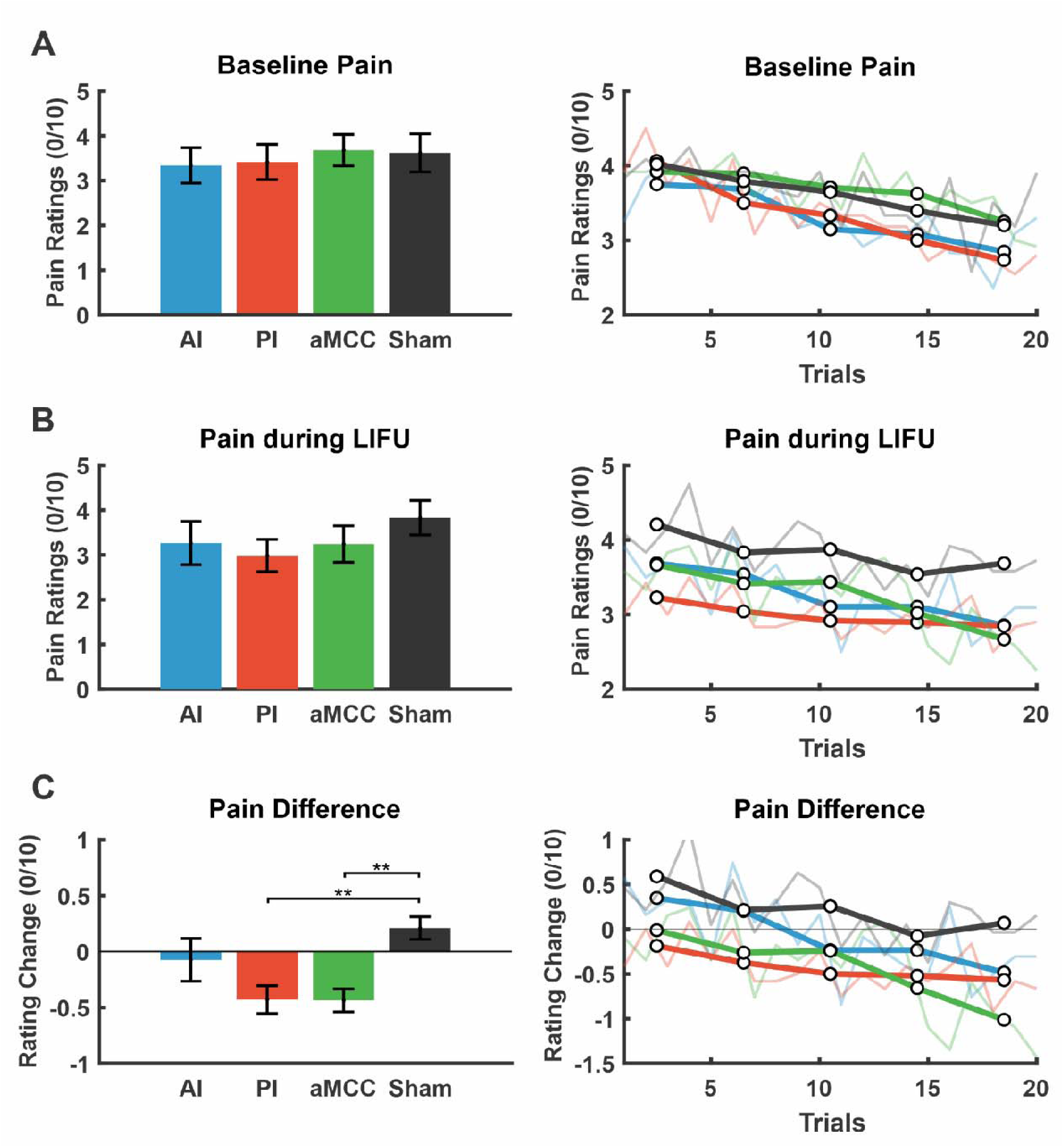
Effect of LIFU on pain ratings. **A.** (Left) Group (N = 12) mean ± SEM baseline pain ratings across LIFU conditions. (Right) Group baseline pain ratings for 20 trials. Thin lines are mean for individual trials. Thick lines represent 4 trial averages. **B.** (Left) Group (N = 12) mean ± SEM pain ratings during LIFU neuromodulation. (Right) Group pain ratings during LIFU for 20 trials. Thin lines are mean for individual trials. Thick lines represent 4 trial averages. **C.** Group (N = 12) mean ± SEM change in pain ratings during LIFU relative to each participant’s same-day baseline rating. A significant effect of condition was observed. Post hoc tests revealed greater pain reduction for PI (**p = 0.001) and aMCC (**p = 0.007) compared with Sham. (Right) Group change in pain rating for 20 trials. Thin lines are mean for individual trials. Thick lines represent 4 trial averages.

### Report of symptoms

Participants completed the Report of Symptoms questionnaire immediately before and 30 minutes after each LIFU visit. Symptoms were rated for presence and severity (mild, moderate, severe). Across all visits, no moderate or severe adverse events were reported. The most frequently reported symptoms were sleepiness, headache, and anxiousness, though reports were mild and not specific to stimulation site or timing (see **Figure S2**). For the Sham condition, the top reported symptoms were headache (n = 2), neck pain (n = 2), and sleepiness (n = 2). In the AI visits, sleepiness (n = 4) was most common, followed by isolated reports of headache (n = 1) and neck pain (n = 1). For the PI condition, participants most often reported sleepiness (n = 3), with occasional headache (n = 1) and neck pain (n = 1). In the aMCC visits, the top symptoms were sleepiness (n = 3), headache (n = 2), and anxiousness (n = 2). To account for pre-existing baseline symptoms, we calculated change scores (pre-post) for each item to determine whether symptoms improved or worsened within each condition. No condition showed strong or consistent worsening (**Figure S2**).

### Auditory masking efficacy

Participants completed a brief experience questionnaire 30 minutes after each visit evaluating whether they could hear or feel the LIFU stimulation and whether they believed they had received stimulation. Responses were collected on a 0-6 Likert scale (0 = “Strongly Disagree,” 1 = “Disagree”, 2 = “Slightly Disagree”, 3 = “Unsure,” 4 = “Slightly Agree”, 5 = “Agree”, 6 = “Strongly Agree”) (**Figure S3**).

For the item “I could hear the LIFU stimulation,” group mean ± SEM ratings were 1.50 ± 0.54, 1.00 ± 0.43, 1.75 ± 0.55, and 1.67 ± 0.56 for AI, PI, aMCC, and Sham, respectively. These values corresponded to subjective ratings ranging from disagree to somewhat disagree that participants could hear the stimulation. The Kruskal-Wallis test found no effect of condition (χ² = 1.20, p = 0.75). For the item “I could feel the LIFU stimulation,” group mean ± SEM ratings were 1.50 ± 0.56, 1.67 ± 0.54, 1.67 ± 0.43, and 1.75 ± 0.58 for AI, PI, aMCC, and Sham, respectively. These ratings likewise corresponded to disagree to somewhat disagree responses, indicating low perceived somatic sensation across all conditions. The Kruskal-Wallis test again found no effect of condition (χ² = 0.63, p = 0.89). For the item “I believe I experienced LIFU stimulation,” group mean ± SEM ratings were 3.00 ± 0.44, 3.17 ± 0.39, 3.08 ± 0.38, and 3.33 ± 0.48 for AI, PI, aMCC, and Sham, respectively. These values clustered around the midpoint of the scale, corresponding to unsure or neutral beliefs regarding whether stimulation was delivered. The Kruskal-Wallis test found no effect of condition (χ² = 0.11, p = 0.99).

## DISCUSSION

This study examined the causal contribution of the AI, PI, and aMCC to sensory gating and subjective ratings of pain intensity during a paired-pulse contact heat evoked potential paradigm in a cohort of healthy participants. At baseline and in the sham condition, participants exhibited intact inhibitory control, as evidenced by robust sensory gating (S2/S1 = 0.57). In contrast, when LIFU was delivered concurrent with the stimuli, LIFU to the PI and aMCC significantly reduced the amplitude of the S2 evoked response relative to sham as well as pain intensity ratings of the S2 stimulus (∼ 0.6 points on a 10-point scale). Contrary to our original hypothesis however, enhanced sensory gating occurred only for LIFU targeting the PI (reduced S2/S1 ratio of ∼28%). Interestingly, there was no evidence that LIFU significantly reduced S1 amplitude compared to Sham. Together, these results suggest that the PI, the earliest cortical recipient of nociceptive input [34], may be a primary site for inhibiting repeated nociceptive signals.

The contribution of the PI to both the subjective experience and inhibitory control of pain is supported by multiple lines of evidence. Functional neuroimaging and source-localized EEG during experimental pain tasks consistently identify robust activity within the PI [69–71] and intracranial recordings demonstrate that, although the PI responds to both innocuous and painful heat, its responses scale only at noxious intensities [37]. Evidence from lesion studies demonstrate that damage of the PI disrupts normal pain perception [72] and direct electrical stimulation of either the PI (and neighboring medial operculum) or the white matter tracts projecting from the ventromedial posterior thalamus into the PI are the only regions of the brain where stimulation can elicit painful sensations [73–75]. Non-human primate tracing studies [35,36], human tractography [76] and resting-state connectivity analysis [77], further reveal the PI as the primary target of lamina I-thalamic afferents and identify dense reciprocal connectivity with the basal ganglia, limbic nuclei, and sensory and salience networks, positioning it as a region capable of regulating ascending nociceptive input. Intracranial recordings across multiple pain-processing regions show that the PI responds earliest to noxious stimulation and drives coordinated activity across widespread brain areas, supporting its role as a cortical entry point for nociceptive signals [34,35,39]. In the present study, LIFU modulation of the PI may have increased gating by shifting the balance between excitation and inhibition towards inhibition within this region. LIFU has been shown to increase GABA and degrees glutamine and glutamate levels in humans [78] and may have transiently elevated GABA within the PI to reduce the magnitude of ascending pain signals. By directly targeting the PI with LIFU during the paired-pulse sensory gating task, these findings extend previous results with causal evidence that the PI directly regulates the flow of ascending nociceptive input and may underlie the sensory gating deficits observed in nociplastic and chronic overlapping pain condition [9,11].

Similar to the PI effects, LIFU to the aMCC reduced subjective ratings of pain intensity as well as the S2 P2P amplitude; however, statistical evidence of sensory gating was lacking for the aMCC [50,79]. Neuroimaging studies consistently identify the aMCC as a core node the salience network [40,80,81], a system that prioritizes behaviorally relevant information. Additional neuroimaging work has also found that activity within the aMCC tracks pain intensity [69,82] and that single unit recordings in humans confirm that neurons within the aMCC selectively respond to and scale at painful stimulation intensities [57]. However, unlike the PI, direct stimulation of the aMCC does not elicit the sensation of pain [57], perhaps due to their distinct thalamic inputs (aMCC: medial dorsal; PI: ventromedial). This distinction suggests that the aMCC may contribute to pain processing differently than the PI. Rather than contributing directly to the generation of pain percepts in the PI, the aMCC may instead top-down modulate the experience of pain via its descending projections to subcortical and brainstem pain modulation centers [83–85]. Consistent with this framework, targeting the aMCC with LIFU reduced both subjective ratings of pain and the S2 response to noxious stimulation. Although aMCC neuromodulation reduced pain and neural responses, it did not alter sensory gating because both the S1 and S2 response amplitudes decreased together (although not significant for S1). As a result, the relative difference between them remained similar, indicating a general dampening of pain processing rather than a selective effect on sensory filtering or gating.

Unlike the S2 response, LIFU neuromodulation did not reduce S1 amplitudes relative to sham for any condition. This finding may reflect fundamental differences in how the two responses are generated. In paired-pulse paradigms, the first stimulus is unexpected and evokes a strong orienting response in addition to nociceptive processing [18], recruiting widespread cortical and subcortical systems common across sensory modalities [24,34,86]. In contrast, the second stimulus is predictable, being time-locked to S1, and is therefore less salient, instead reflecting more modality-specific nociceptive processing within relatively localized cortical circuitry [56]. Given the high spatial specificity of LIFU, its effects would be expected to emerge more clearly in localized nociceptive responses such as S2 than in broadly distributed salience-related responses such as S1. Consistent with this framework, targeting the PI, a region specialized for nociceptive but not salience processing, preferentially modulated the S2 response while leaving S1 largely unchanged. In contrast, the aMCC, which is embedded within the salience network, would be expected to influence the more globally distributed S1 response; however, modulation of this salience-driven component was not detected in the present study. Notably, aMCC targeting also reduced S2 nociception-weighted amplitudes, consistent with its role in early thalamocortical processing [34].

LIFU neuromodulation of the AI did not significantly change pain ratings, S1 or S2 CHEP amplitudes, or sensory gating. This result was unexpected, as we initially predicted that AI neuromodulation would be similar to aMCC effects due to their shared involvement in the salience network [40]. However, the absence of AI effects may reflect fundamental differences in how the AI and aMCC contribute to pain processing. The aMCC receives direct nociceptive input via the spinothalamic tract through the mediodorsal thalamus [35]; is known to encode pain intensity [57], and is positioned to shape ascending nociceptive signals through its broader connections with motor-control regions [36,42]. In contrast, the AI does not receive direct nociceptive input via the spinothalamic tract through thalamic nuclei [39]. Instead, pain-related information looks to reach the AI indirectly through the PI [34], where it is generally thought to be combined with affective and autonomic signals, thereby influencing both the emotional experience of and the body’s physiological response to pain [38,87]. Supporting this idea, our recent study targeting the AI also found no change in pain intensity but did instead demonstrate shifts in heart rate variability [54]. Together, these findings indicate that the PI and aMCC are more tightly linked to early sensory processing and inhibitory control, whereas the AI may play a more integrative role that does not strongly influence gating mechanisms.

## LIMITATIONS

This study included only healthy adults, who exhibited intact sensory gating at baseline, with S2/S1 ratios ranging between 0.41 and 0.75 across participants. Because chronic pain is associated with altered sensory gating and changes in insula-cingulate connectivity [88,89], the neuromodulatory effects observed here may not generalize to clinical pain populations. These results should therefore be interpreted as characterizing LIFU effects for healthy pain processing. Additionally, this study only recorded subjective pain intensity and did not assess the emotional or unpleasant aspects of pain, which may be more sensitive to AI neuromodulation and could reveal effects that were not detectable with intensity ratings alone. Finally, the timing of LIFU delivery should be considered when interpreting the different effects on S1 and S2. LIFU was applied across both responses, resulting in a longer exposure before the S2 stimulus presentation. Because longer LIFU delivery is known to produce stronger effects [64,90], this timing may have favored modulation of S2. As a result, the greater reduction in S2 amplitude may partly reflect increased exposure rather than differences between the responses themselves. Future work should consider applying offline neuromodulation paradigms to avoid this concern.

## CONCLUSIONS

LIFU to the PI and aMCC reduced subjective pain intensity ratings and the S2 P2P amplitude, but only PI neuromodulation enhanced sensory gating. These results show that sensory gating can be increased in healthy adults and point to the PI as a key site involved in regulating repeated nociceptive signals. Future studies should test whether modulating these regions in clinical populations with impaired gating, such as those with nociplastic pain, can help restore inhibitory control and reduce pain hypersensitivity.

## Data Availability

All data produced in the present study are available upon reasonable request to the authors.

## ACKNOWLEDGEMENTS

This work was funded in part by grants awarded to W.L. from the National Institute for Health (NIH) 1R21AT012247-01. The authors would like to thank Jessica Florig, Brighton Payne, John Hanrahan, and Kathryn Painchaud for their help with data collection and analysis. Data availability statement: Data are available upon reasonable request. Please contact the corresponding author.

## Supplemental Figures

**Figure S1.**
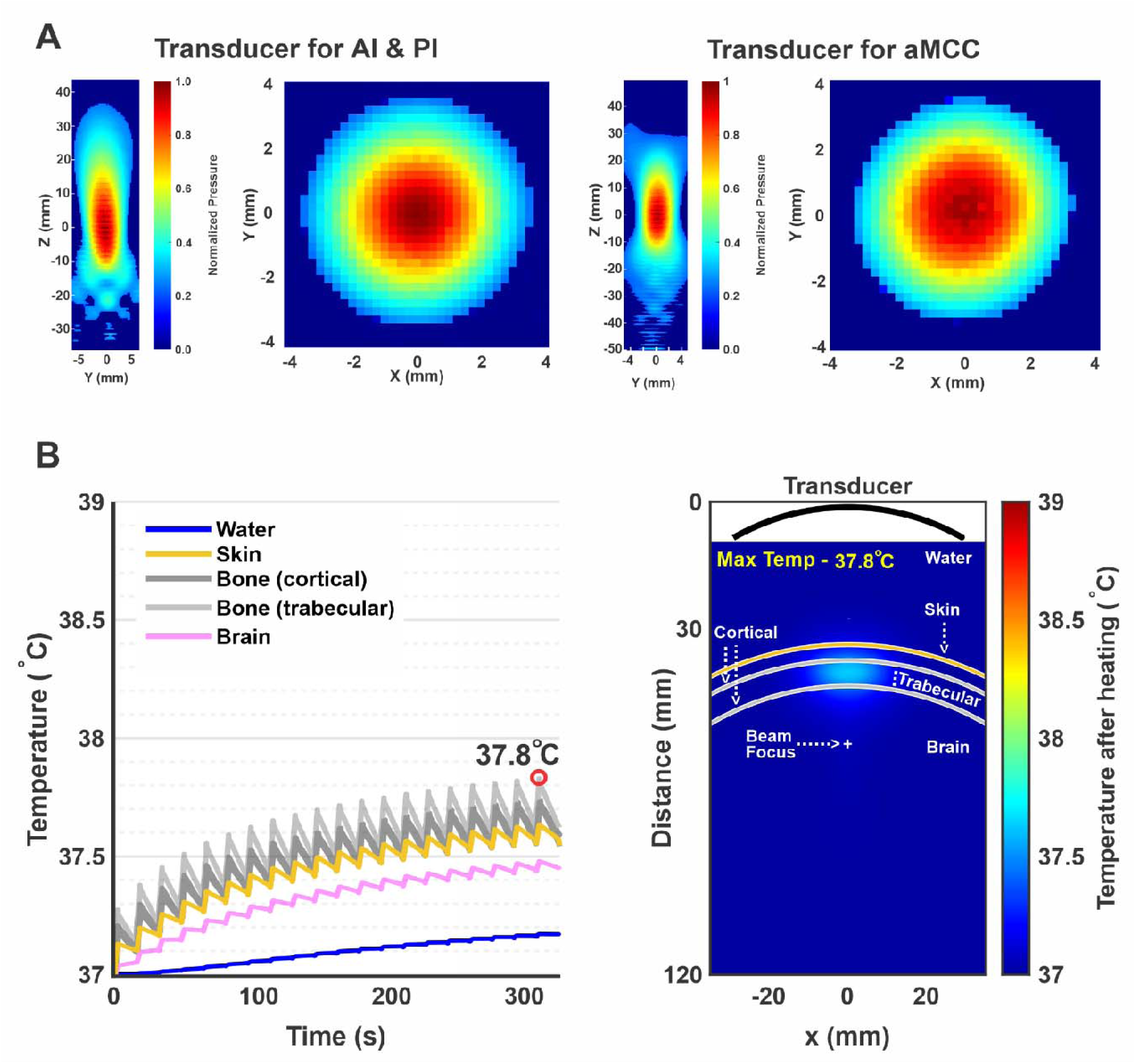
Acoustic beam profiles and thermal safety of LIFU stimulation. **A.** Pseudocolor YZ and XY plane empirical measurements obtained in an acoustic test tank showing the pressure field of the transducer used to target the anterior insula (AI) and posterior insula (PI) (left) and the anterior mid-cingulate cortex (aMCC) (right). **B.** Thermal modeling results using a three-layer skull model (cortical-trabecular-cortical) under conservative stimulation conditions (30% duty cycle, 2.5-s pulse duration, 10 Hz PRF, 750 kPa extracranial pressure, 20 repetitions, 14-s interstimulus interval). (Left) Temperature rise over time in water, skin, cortical bone, trabecular bone, and brain tissue. Red dot denotes highest estimated temperature. (Right) Spatial temperature distribution showing the beam focus and skull layers. The maximum temperature occurred in cortical bone (37.8 °C; +0.83 °C above baseline), while peak brain temperature reached 37.5 °C. All simulated temperature increases remained well below the 2 °C safety limit recommended by current guidelines.

**Figure S2.**
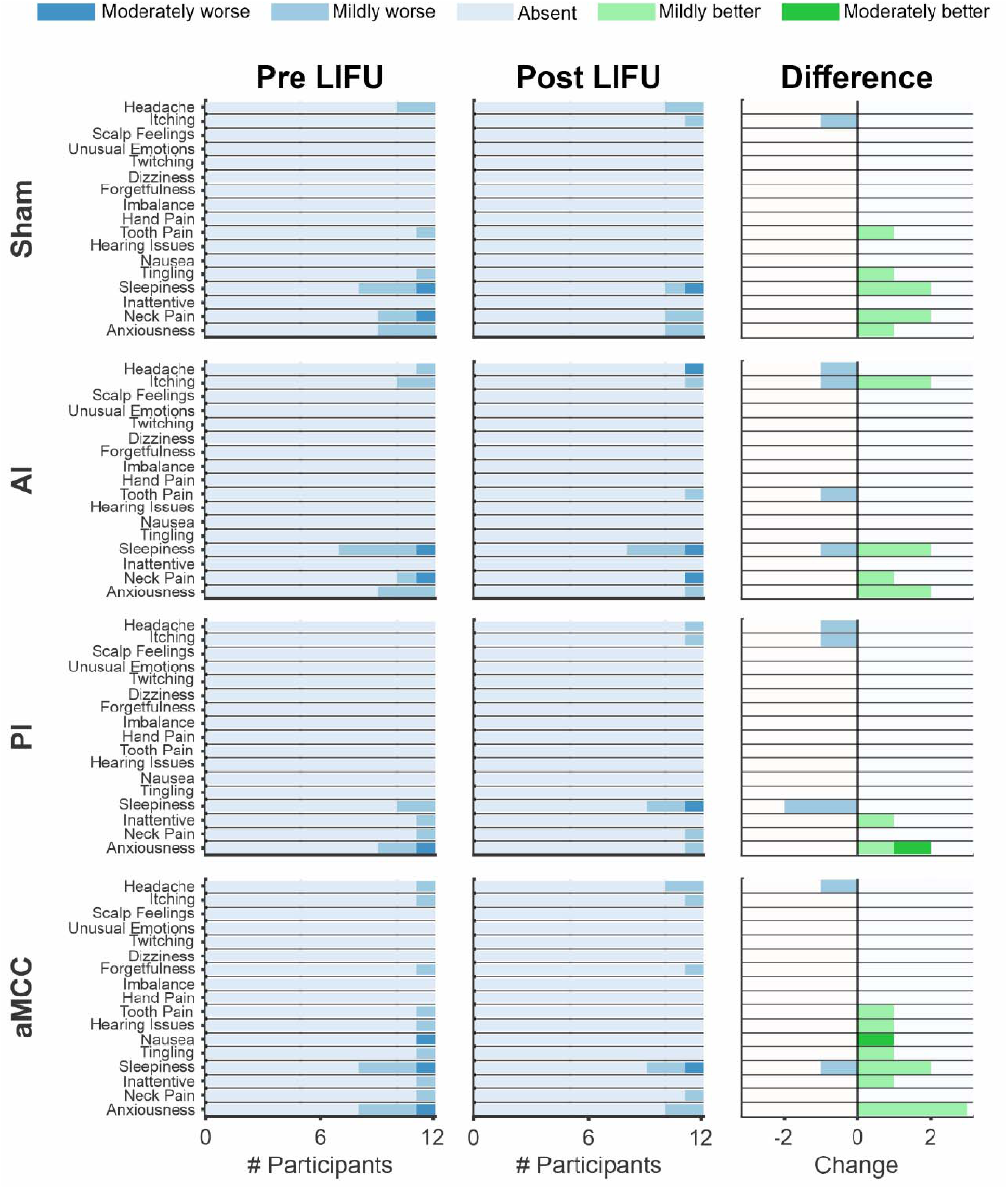
Self-reported symptoms before and after each session (N = 12). Participants completed a report-of-symptoms questionnaire before testing and again 30 minutes after each session. Rows show individual symptoms for each condition (Sham, AI, PI, aMCC), and columns display responses before LIFU, after LIFU, and the change between the two. Bar lengths indicate the number of participants reporting each level of symptom severity. In the difference panels, green reflects improvement and blue reflects worsening of symptoms; “absent” responses are omitted to highlight directional shifts. Overall, symptom profiles were similar before and after sessions, with no consistent pattern of worsening following LIFU.

**Figure S3.**
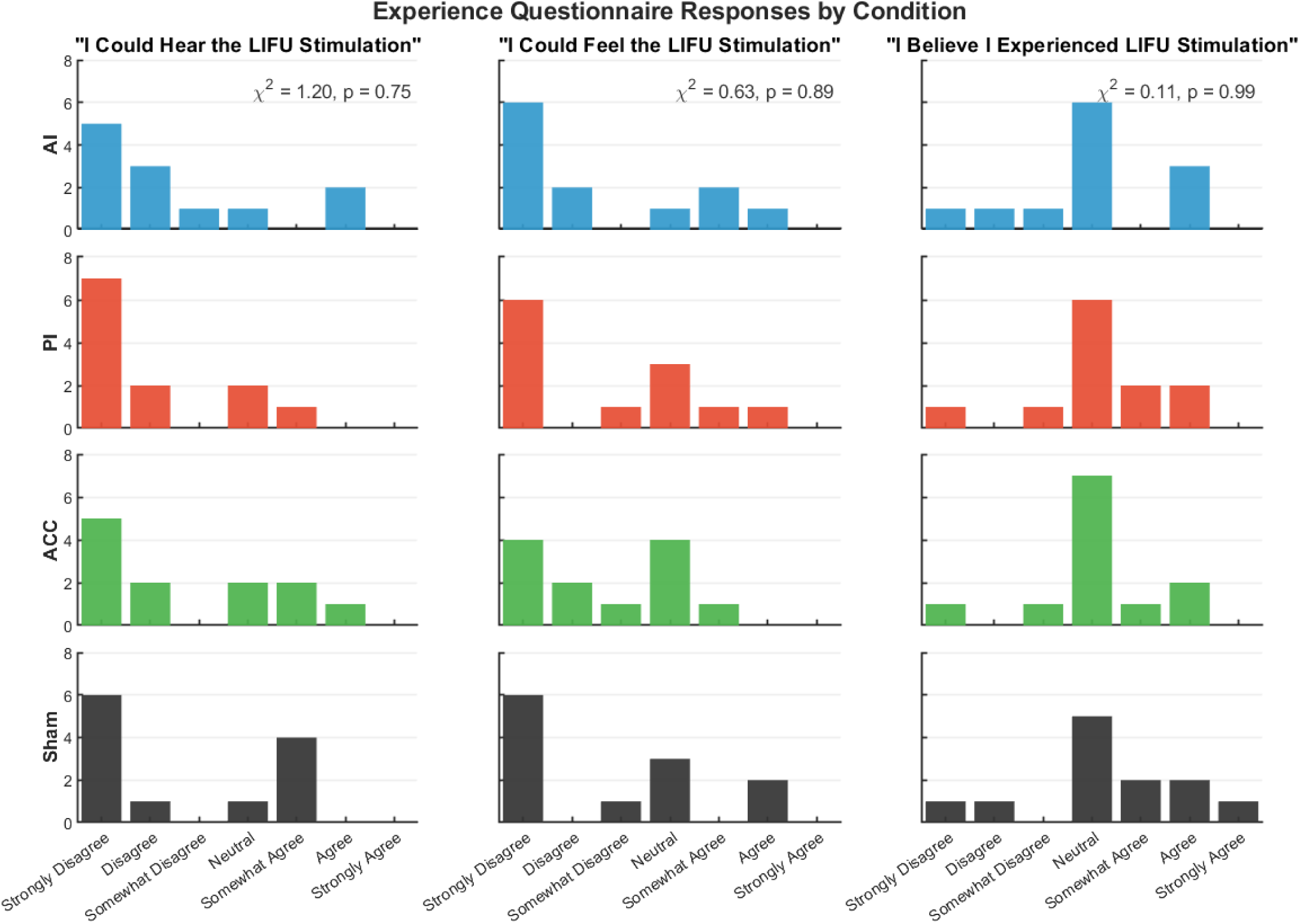
Experience questionnaire responses across study conditions (N = 12). Distributions of participant ratings for three items: “I could hear the LIFU stimulation,” “I could feel the LIFU stimulation,” and “I believe I experienced LIFU stimulation.” Each row shows a stimulation condition (AI, PI, aMCC, Sham), and each column corresponds to one question. Bars indicate the number of participants selecting each response on a 7-point Likert scale (strongly disagree to strongly agree). No differences were observed between conditions for any item (Kruskal-Wallis tests shown in the top row), indicating that participants were unable to reliably detect whether stimulation was delivered.

## Notes

### Competing Interest Statement

The authors have declared no competing interest.

### Clinical Trial

NCT05145426

### Author Declarations

All experimental procedures were deemed ethical and approved by the Virginia Tech Institutional Review Board (VT-IRB #21-796).

## References

[1] Venables PH. INPUT DYSFUNCTION IN SCHIZOPHRENIA. Prog Exp Pers Res 1964;72:1–47.

[2] Cromwell HC, Mears RP, Wan L, Boutros NN. Sensory Gating: A Translational Effort from Basic to Clinical Science. Clin EEG Neurosci 2008;39:69–72. 10.1177/155005940803900209.

[3] Glaser EM, Griffin JP. Influence of the cerebral cortex on habituation. The Journal of Physiology 1962;160:429–45. 10.1113/jphysiol.1962.sp006857.

[4] Adler LE, Pachtman E, Franks RD, Pecevich M, Waldo MC, Freedman R. Neurophysiological evidence for a defect in neuronal mechanisms involved in sensory gating in schizophrenia. Biol Psychiatry 1982;17:639–54.

[5] Lijffijt M, Lane SD, Meier SL, Boutros NN, Burroughs S, Steinberg JL, et al. P50, N100, and P200 sensory gating: Relationships with behavioral inhibition, attention, and working memory. Psychophysiology 2009;46:1059–68. 10.1111/j.1469-8986.2009.00845.x.

[6] Harris JD. Habituatory response decrement in the intact organism. Psychological Bulletin 1943;40:385–422. 10.1037/h0053918.

[7] Thompson RF, Spencer WA. Habituation: A model phenomenon for the study of neuronal substrates of behavior. Psychological Review 1966;73:16–43. 10.1037/h0022681.

[8] Morse K, Vander Werff KR. Cortical Auditory Evoked Potential Indices of Impaired Sensory Gating in People With Chronic Tinnitus. Ear and Hearing 2024;45:730. 10.1097/AUD.0000000000001463.

[9] de Tommaso M, Federici A, Santostasi R, Calabrese R, Vecchio E, Lapadula G, et al. Laser-Evoked Potentials Habituation in Fibromyalgia. The Journal of Pain 2011;12:116–24. 10.1016/j.jpain.2010.06.004.

[10] Lim M, Roosink M, Kim JS, Kim DJ, Kim HW, Lee EB, et al. Disinhibition of the primary somatosensory cortex in patients with fibromyalgia. PAIN 2015;156:666. 10.1097/j.pain.0000000000000096.

[11] Montoya P, Sitges C, García-Herrera M, Rodríguez-Cotes A, Izquierdo R, Truyols M, et al. Reduced brain habituation to somatosensory stimulation in patients with fibromyalgia. Arthritis Rheum 2006;54:1995–2003. 10.1002/art.21910.

[12] Lenz M, Höffken O, Stude P, Lissek S, Schwenkreis P, Reinersmann A, et al. Bilateral somatosensory cortex disinhibition in complex regional pain syndrome type I. Neurology 2011;77:1096–101. 10.1212/WNL.0b013e31822e1436.

[13] Fruhstorfer H, Soveri P, Järvilehto T. Short-term habituation of the auditory evoked response in man. Electroencephalography and Clinical Neurophysiology 1970;28:153–61. 10.1016/0013-4694(70)90183-5.

[14] Budd TW, Barry RJ, Gordon E, Rennie C, Michie PT. Decrement of the N1 auditory event-related potential with stimulus repetition: habituation vs. refractoriness. International Journal of Psychophysiology 1998;31:51–68. 10.1016/S0167-8760(98)00040-3.

[15] Hari R, Kaila K, Katila T, Tuomisto T, Varpula T. Interstimulus interval dependence of the auditory vertex response and its magnetic counterpart: Implications for their neural generation. Electroencephalography and Clinical Neurophysiology 1982;54:561–9. 10.1016/0013-4694(82)90041-4.

[16] Järvilehto T, Hari R, Sams M. Effect of stimulus repetition on negative sustained potentials elicited by auditory and visual stimuli in the human EEG. Biological Psychology 1978;7:1–12. 10.1016/0301-0511(78)90038-8.

[17] Siegel C, Waldo M, Mizner G, Adler LE, Freedman R. Deficits in Sensory Gating in Schizophrenic Patients and Their Relatives: Evidence Obtained With Auditory Evoked Responses. Arch Gen Psychiatry 1984;41:607–12. 10.1001/archpsyc.1984.01790170081009.

[18] Legrain V, Iannetti GD, Plaghki L, Mouraux A. The pain matrix reloaded: A salience detection system for the body. Progress in Neurobiology 2011;93:111–24. 10.1016/j.pneurobio.2010.10.005.

[19] Choi W, Lim M, Kim JS, Chung CK. Habituation deficit of auditory N100m in patients with fibromyalgia. Eur J Pain 2016;20:1634–43. 10.1002/ejp.883.

[20] Chen I-A, Hung SW, Chen Y-H, Lim S-N, Tsai Y-T, Hsiao C-L, et al. Contact Heat Evoked Potentials in Normal Subjects 2006;15.

[21] Granovsky Y, Anand P, Nakae A, Nascimento O, Smith B, Sprecher E, et al. Normative data for Aδ contact heat evoked potentials in adult population: a multicenter study. PAIN 2016;157:1156. 10.1097/j.pain.0000000000000495.

[22] Granovsky Y, Granot M, Nir R-R, Yarnitsky D. Objective Correlate of Subjective Pain Perception by Contact Heat-Evoked Potentials. The Journal of Pain 2008;9:53–63. 10.1016/j.jpain.2007.08.010.

[23] Mouraux A, Diukova A, Lee MC, Wise RG, Iannetti GD. A multisensory investigation of the functional significance of the “pain matrix.” NeuroImage 2011;54:2237–49. 10.1016/j.neuroimage.2010.09.084.

[24] Somervail R, Perovic S, Bufacchi RJ, Caminiti R, Iannetti GD. A two-system theory of sensory-evoked brain responses. Brain 2025:awaf402. 10.1093/brain/awaf402.

[25] Kaplan CM, Kelleher E, Irani A, Schrepf A, Clauw DJ, Harte SE. Deciphering nociplastic pain: clinical features, risk factors and potential mechanisms. Nat Rev Neurol 2024;20:347–63. 10.1038/s41582-024-00966-8.

[26] Nahman-Averbuch H, Martucci KT, Granovsky Y, Weissman-Fogel I, Yarnitsky D, Coghill RC. Distinct brain mechanisms support spatial vs temporal filtering of nociceptive information. PAIN 2014;155:2491. 10.1016/j.pain.2014.07.008.

[27] Piché M, Arsenault M, Rainville P. Cerebral and Cerebrospinal Processes Underlying Counterirritation Analgesia. J Neurosci 2009;29:14236–46. 10.1523/JNEUROSCI.2341-09.2009.

[28] Youssef AM, Macefield VG, Henderson LA. Cortical influences on brainstem circuitry responsible for conditioned pain modulation in humans. Hum Brain Mapp 2016;37:2630–44. 10.1002/hbm.23199.

[29] Emmert K, Breimhorst M, Bauermann T, Birklein F, Van De Ville D, Haller S. Comparison of anterior cingulate vs. insular cortex as targets for real-time fMRI regulation during pain stimulation. Front Behav Neurosci 2014;8. 10.3389/fnbeh.2014.00350.

[30] Mosch B, Hagena V, Herpertz S, Ruttorf M, Diers M. Neural correlates of control over pain in fibromyalgia patients. Neuroimage Clin 2023;37:103355. 10.1016/j.nicl.2023.103355.

[31] Starr CJ, Sawaki L, Wittenberg GF, Burdette JH, Oshiro Y, Quevedo AS, et al. Roles of the Insular Cortex in the Modulation of Pain: Insights from Brain Lesions. Journal of Neuroscience 2009;29:2684–94. 10.1523/JNEUROSCI.5173-08.2009.

[32] Brooks JCW, Tracey I. The insula: A multidimensional integration site for pain. Pain 2007;128:1–2. 10.1016/j.pain.2006.12.025.

[33] Cauda F, D’Agata F, Sacco K, Duca S, Geminiani G, Vercelli A. Functional connectivity of the insula in the resting brain. Neuroimage 2011;55:8–23. 10.1016/j.neuroimage.2010.11.049.

[34] Bastuji H, Frot M, Perchet C, Magnin M, Garcia-Larrea L. Pain networks from the inside: Spatiotemporal analysis of brain responses leading from nociception to conscious perception. Hum Brain Mapp 2016;37:4301–15. 10.1002/hbm.23310.

[35] Craig AD. Pain mechanisms: Labeled lines versus convergence in central processing. Annual Review of Neuroscience 2003;26:1–30.

[36] Dum RP, Levinthal DJ, Strick PL. The Spinothalamic System Targets Motor and Sensory Areas in the Cerebral Cortex of Monkeys. J Neurosci 2009;29:14223–35. 10.1523/JNEUROSCI.3398-09.2009.

[37] Frot M, Magnin M, Mauguière F, Garcia-Larrea L. Human SII and Posterior Insula Differently Encode Thermal Laser Stimuli. Cerebral Cortex 2007;17:610–20. 10.1093/cercor/bhk007.

[38] Uddin LQ. Salience processing and insular cortical function and dysfunction | Nature Reviews Neuroscience. Nature Reviews Neuroscience 2015;16:55–61. 10.1038/nrn3857.

[39] Bastuji H, Frot M, Perchet C, Hagiwara K, Garcia-Larrea L. Convergence of sensory and limbic noxious input into the anterior insula and the emergence of pain from nociception. Scientific Reports 2018;8:13360. 10.1038/s41598-018-31781-z.

[40] Seeley WW, Menon V, Schatzberg AF, Keller J, Glover GH, Kenna H, et al. Dissociable Intrinsic Connectivity Networks for Salience Processing and Executive Control. Journal of Neuroscience 2007;27:2349–56. 10.1523/JNEUROSCI.5587-06.2007.

[41] Fiddick L. There is more than the amygdala: Potential threat assessment in the cingulate cortex. Neuroscience & Biobehavioral Reviews 2011;35:1007–18. 10.1016/j.neubiorev.2010.09.014.

[42] Dosenbach NUF, Raichle ME, Gordon EM. The brain’s action-mode network. Nat Rev Neurosci 2025;26:158–68. 10.1038/s41583-024-00895-x.

[43] Kuner R, Kuner T. Cellular Circuits in the Brain and Their Modulation in Acute and Chronic Pain. Physiological Reviews 2021;101:213–58. 10.1152/physrev.00040.2019.

[44] Gracely RH, Petzke F, Wolf JM, Clauw DJ. Functional magnetic resonance imaging evidence of augmented pain processing in fibromyalgia. Arthritis & Rheumatism 2002;46:1333–43. 10.1002/art.10225.

[45] López-Solà M, Woo C-W, Pujol J, Deus J, Harrison BJ, Monfort J, et al. Towards a neurophysiological signature for fibromyalgia. PAIN 2017;158:34. 10.1097/j.pain.0000000000000707.

[46] Harris RE, Sundgren PC, Craig AD (Bud), Kirshenbaum E, Sen A, Napadow V, et al. Elevated Insular Glutamate (Glu) in Fibromyalgia (FM) is Associated with Experimental Pain. Arthritis Rheum 2009;60:3146–52. 10.1002/art.24849.

[47] Harte SE, Ichesco E, Hampson JP, Peltier SJ, Schmidt-Wilcke T, Clauw DJ, et al. Pharmacologic attenuation of cross-modal sensory augmentation within the chronic pain insula. PAIN 2016;157:1933. 10.1097/j.pain.0000000000000593.

[48] Hubbard CS, Lazaridou A, Cahalan CM, Kim J, Edwards RR, Napadow V, et al. Aberrant salience? Brain hyperactivation in response to pain onset and offset in fibromyalgia. Arthritis Rheumatol 2020;72:1203–13. 10.1002/art.41220.

[49] Spagnolo PA, Wang H, Srivanitchapoom P, Schwandt M, Heilig M, Hallett M. Lack of Target Engagement Following Low-Frequency Deep Transcranial Magnetic Stimulation of the Anterior Insula. Neuromodulation: Technology at the Neural Interface 2019;22:877–83. 10.1111/ner.12875.

[50] Strohman A, Legon W. Neuromodulation of the Cingulate Cortex for Pain. Neuroscientist 2025:10738584251337652. 10.1177/10738584251337652.

[51] Legon W, Sato TF, Opitz A, Mueller J, Barbour A, Williams A, et al. Transcranial focused ultrasound modulates the activity of primary somatosensory cortex in humans. Nat Neurosci 2014;17:322–9. 10.1038/nn.3620.

[52] Legon W, Strohman A. Low-intensity focused ultrasound for human neuromodulation. Nat Rev Methods Primers 2024;4:91. 10.1038/s43586-024-00368-6.

[53] Legon W, Bansal P, Tyshynsky R, Ai L, Mueller JK. Transcranial focused ultrasound neuromodulation of the human primary motor cortex. Sci Rep 2018;8:10007. 10.1038/s41598-018-28320-1.

[54] Legon W, Strohman A, In A, Payne B. Noninvasive neuromodulation of subregions of the human insula differentially affect pain processing and heart-rate variability: a within-subjects pseudo-randomized trial. PAIN 2024:10.1097/j.pain.0000000000003171. https://doi.org/10.1097/j.pain.0000000000003171.

[55] Strohman A, Payne B, In A, Stebbins K, Legon W. Low-intensity focused ultrasound to the human dorsal anterior cingulate attenuates acute pain perception and autonomic responses. J Neurosci 2024. 10.1523/JNEUROSCI.1011-23.2023.

[56] Ronga I, Valentini E, Mouraux A, Iannetti GD. Novelty is not enough: laser-evoked potentials are determined by stimulus saliency, not absolute novelty. Journal of Neurophysiology 2013;109:692–701. 10.1152/jn.00464.2012.

[57] Hutchison WD, Davis KD, Lozano AM, Tasker RR, Dostrovsky JO. Pain-related neurons in the human cingulate cortex. Nat Neurosci 1999;2:403–5. 10.1038/8065.

[58] Rossi S, Hallett M, Rossini PM, Pascual-Leone A. Safety, ethical considerations, and application guidelines for the use of transcranial magnetic stimulation in clinical practice and research. Clinical Neurophysiology 2009;120:2008–39. 10.1016/j.clinph.2009.08.016.

[59] Legon W, Adams S, Bansal P, Patel PD, Hobbs L, Ai L, et al. A retrospective qualitative report of symptoms and safety from transcranial focused ultrasound for neuromodulation in humans. Sci Rep 2020;10:5573. 10.1038/s41598-020-62265-8.

[60] Kapoor A, Strohman A, Ni Y, Isaac G, Raymond J, Legon W. Acoustic Coupling for Double-Blind Human Low-Intensity Focused Ultrasound Neuromodulation 2025:2025.10.02.680055. 10.1101/2025.10.02.680055.

[61] Liang W, Guo H, Mittelstein DR, Shapiro MG, Shimojo S, Shehata MH. Auditory Mondrian masks the airborne-auditory artifact of focused ultrasound stimulation in humans. Brain Stimulation: Basic, Translational, and Clinical Research in Neuromodulation 2023;0. 10.1016/j.brs.2023.03.002.

[62] Jensen MP, Karoly P. Self-report scales and procedures for assessing pain in adults. Handbook of pain assessment, 2nd ed, New York, NY, US: The Guilford Press; 2001, p. 15–34.

[63] Strohman A, In A, Stebbins K, Legon W. Evaluation of a Novel Acoustic Coupling Medium for Human Low-Intensity Focused Ultrasound Neuromodulation Applications. Ultrasound in Medicine & Biology 2023. 10.1016/j.ultrasmedbio.2023.02.003.

[64] Ennasr A, Isaac G, Strohman A, Legon W. Examination of the interaction of parameters for low-intensity focused ultrasound of the human motor cortex. Brain Stimulation 2024. 10.1016/j.brs.2024.11.005.

[65] Treeby BE, Cox BT. k-Wave: MATLAB toolbox for the simulation and reconstruction of photoacoustic wave fields. J Biomed Opt 2010;15:021314. 10.1117/1.3360308.

[66] Legon W, Ai L, Bansal P, Mueller JK. Neuromodulation with single-element transcranial focused ultrasound in human thalamus. Hum Brain Mapp 2018;39:1995–2006. 10.1002/hbm.23981.

[67] Aubry J-F, Bates O, Boehm C, Butts Pauly K, Christensen D, Cueto C, et al. Benchmark problems for transcranial ultrasound simulation: Intercomparison of compressional wave modelsa). The Journal of the Acoustical Society of America 2022;152:1003–19. 10.1121/10.0013426.

[68] Saito Y, Tsuchiya T, Endoh N. Numerical Analysis of Temperature Rise in Tissue Using Electronically Focused Ultrasound. Jjap 2006;45:4693. 10.1143/JJAP.45.4693.

[69] Wager TD, Atlas LY, Lindquist MA, Roy M, Woo C-W, Kross E. An fMRI-Based Neurologic Signature of Physical Pain. N Engl J Med 2013;368:1388–97. 10.1056/NEJMoa1204471.

[70] Valeriani M, Le Pera D, Niddam D, Chen ACN, Arendt-Nielsen L. Dipolar modelling of the scalp evoked potentials to painful contact heat stimulation of the human skin. Neuroscience Letters 2002;318:44–8. 10.1016/S0304-3940(01)02466-1.

[71] Segerdahl AR, Mezue M, Okell TW, Farrar JT, Tracey I. The dorsal posterior insula subserves a fundamental role in human pain. Nat Neurosci 2015;18:499–500. 10.1038/nn.3969.

[72] Greenspan JD, Lee RR, Lenz FA. Pain sensitivity alterations as a function of lesion location in the parasylvian cortex. Pain 1999;81:273–82. 10.1016/S0304-3959(99)00021-4.

[73] Ostrowsky K, Magnin M, Ryvlin P, Isnard J, Guenot M, Mauguière F. Representation of Pain and Somatic Sensation in the Human Insula: a Study of Responses to Direct Electrical Cortical Stimulation. Cereb Cortex 2002;12:376–85. 10.1093/cercor/12.4.376.

[74] Mandonnet V, Obaid S, Descoteaux M, St-Onge E, Devaux B, Levé C, et al. Electrostimulation of the white matter of the posterior insula and medial operculum: perception of vibrations, heat, and pain. Pain 2024;165:565–72. 10.1097/j.pain.0000000000003069.

[75] Mazzola L, Isnard J, Peyron R, Mauguière F. Stimulation of the human cortex and the experience of pain: Wilder Penfield’s observations revisited. Brain 2012;135:631–40. 10.1093/brain/awr265.

[76] Ghaziri J, Tucholka A, Girard G, Houde J-C, Boucher O, Gilbert G, et al. The Corticocortical Structural Connectivity of the Human Insula. Cerebral Cortex 2017;27:1216–28. 10.1093/cercor/bhv308.

[77] Wiech K, Jbabdi S, Lin CS, Andersson J, Tracey I. Differential structural and resting state connectivity between insular subdivisions and other pain-related brain regions. Pain 2014;155:2047–55. 10.1016/j.pain.2014.07.009.

[78] Zhang T, Guo B, Zuo Z, Long X, Hu S, Li S, et al. Excitatory-inhibitory modulation of transcranial focus ultrasound stimulation on human motor cortex. CNS Neuroscience & Therapeutics 2023;29:3829–41. 10.1111/cns.14303.

[79] Wager TD, Atlas LY, Botvinick MM, Chang LJ, Coghill RC, Davis KD, et al. Pain in the ACC? Proceedings of the National Academy of Sciences 2016;113:E2474–5. 10.1073/pnas.1600282113.

[80] Dosenbach NUF, Fair DA, Miezin FM, Cohen AL, Wenger KK, Dosenbach RAT, et al. Distinct brain networks for adaptive and stable task control in humans. Proceedings of the National Academy of Sciences 2007;104:11073–8. 10.1073/pnas.0704320104.

[81] Menon V, Uddin LQ. Saliency, switching, attention and control: a network model of insula function. Brain Struct Funct 2010;214:655–67. 10.1007/s00429-010-0262-0.

[82] Xu A, Larsen B, Baller EB, Scott JC, Sharma V, Adebimpe A, et al. Convergent neural representations of experimentally-induced acute pain in healthy volunteers: A large-scale fMRI meta-analysis. Neuroscience & Biobehavioral Reviews 2020;112:300–23. 10.1016/j.neubiorev.2020.01.004.

[83] Beissner F, Meissner K, Bär K-J, Napadow V. The Autonomic Brain: An Activation Likelihood Estimation Meta-Analysis for Central Processing of Autonomic Function. J Neurosci 2013;33:10503–11. 10.1523/JNEUROSCI.1103-13.2013.

[84] Kleckner IR, Zhang J, Touroutoglou A, Chanes L, Xia C, Simmons WK, et al. Evidence for a large-scale brain system supporting allostasis and interoception in humans. Nat Hum Behav 2017;1:1–14. 10.1038/s41562-017-0069.

[85] Chen T, Taniguchi W, Chen Q-Y, Tozaki-Saitoh H, Song Q, Liu R-H, et al. Top-down descending facilitation of spinal sensory excitatory transmission from the anterior cingulate cortex. Nat Commun 2018;9:1886. 10.1038/s41467-018-04309-2.

[86] Mouraux A, Iannetti GD. Nociceptive Laser-Evoked Brain Potentials Do Not Reflect Nociceptive-Specific Neural Activity. Journal of Neurophysiology 2009;101:3258–69. 10.1152/jn.91181.2008.

[87] Frot M, Faillenot I, Mauguière F. Processing of nociceptive input from posterior to anterior insula in humans. Human Brain Mapping 2014;35:5486–99. 10.1002/hbm.22565.

[88] Napadow V, LaCount L, Park K, As-Sanie S, Clauw DJ, Harris RE. Intrinsic brain connectivity in fibromyalgia is associated with chronic pain intensity. Arthritis Rheum 2010;62:2545–55. 10.1002/art.27497.

[89] Pujol J, López-Solà M, Ortiz H, Vilanova JC, Harrison BJ, Yücel M, et al. Mapping Brain Response to Pain in Fibromyalgia Patients Using Temporal Analysis of fMRI. PLOS ONE 2009;4:e5224. 10.1371/journal.pone.0005224.

[90] Zeng K, Li Z, Xia X, Wang Z, Darmani G, Li X, et al. Effects of different sonication parameters of theta burst transcranial ultrasound stimulation on human motor cortex. Brain Stimulation 2024;17:258–68. 10.1016/j.brs.2024.03.001.

